# Impact of COVID-19 on the Psychological Well-Being and Turnover Intentions of Frontline Nurses in the Community: A Cross-Sectional Study in the Philippines

**DOI:** 10.1101/2020.08.05.20167411

**Authors:** Janet Alexis A. De los Santos, Leodoro J. Labrague

**Affiliations:** Visayas State University Philippines; Sultan Qaboos University, Oman

**Keywords:** COVID-19 pandemic, fear, turnover intentions, psychological well-being, nursing

## Abstract

**Purpose:** This study aimed to assess fear of COVID-19 among nurses in a community setting.

**Methods:** This study employed a cross-sectional design using self-report questionnaires.

**Findings:** Results revealed that nurses display moderate to high fear of COVID-19 and that the female gender is correlated to fear of the virus. Moreover, the nurses’ fear influences their psychological distress and organizational and professional turnover intentions.

**Conclusion:** Fear of COVID-19 is universal among nurses. There is a need to assess the factors associated with the fear to better address the nurses’ psychological well-being and to avoid turnover intentions.

## INTRODUCTION

The novel coronavirus virus, now known as COVID-19, first erupted in December 2019 in Chinese territory, particularly in Wuhan, China. The impact of this dreadful disease on jobs, the economy, and the personal lives of people globally is unprecedented. This novel virus’s exponential effect has caused large-scale closing of economies, loss of employment, uncomfortable living adjustments, and the untimely death of loved ones. The highly infectious respiratory disease has reached more than 200 countries, hence its pandemic status. As of July 19, 2020, there have been 14,043,176 confirmed cases of the virus, resulting in the loss of 597,583 lives across the globe (WHO, 2020). An estimated 230,000 of these cases were healthcare workers, and 600 of those were nurses who contracted the disease while performing their duties caring for those afflicted with COVID-19 (International Council of Nurses, 2020).

In the Philippines, the nationwide cases have reached about 67,457, with 43,160 of those active, and 1,831 have died as of the latest count (DOH, 2020). To aid the economy, the Philippine government has shifted its propaganda from ‘stay at home’ and strict quarantine protocols to meticulous handwashing, physical distancing, and wearing of a mask, especially when going out in public. The Philippines’ Department of Health has firmly advocated compliance to at least the minimum health standards and protocols to avert the collapse of health institutes and to assist the health workforce responding to COVID-19 cases in the country. To boost the morale of the frontline health workers, the Duterte administration initiated monetary remuneration for doctors and nurses who incur the virus in their line of work. As of June 22, 2020, there are 3,122 health workers in the country inflicted with the virus, composed mainly of nurses (Esguerra, 2020).

With the increasing death toll of health workers caring for patients with COVID-19 are the psychological challenges. The uncertainty and discomfort in this new normal may pose a further threat to healthcare professionals’ work outcomes and psychological wellbeing. Nurses are placed in stressful situations, fulfilling their roles on the front line while risking their lives to save others.

### Review of Related Literature

Recent literature has established the ill effects of stress on the nurses’ psychological well-being and work outcomes (Falguera et al., 2020; Faremi et al., 2019; Vivian et al., 2019). Stress is generally sourced from situations that a person has no control over, such as a pandemic. Currently, there is a surge of studies on how the COVID-19 pandemic has caused much stress to the various healthcare systems across the globe. It has compromised the workforce, particularly nurses. In fact, among the healthcare workers, nurses are found to be the most anxious and stressed in caring for and treating patients infected with the COVID-19 virus (Mo et al., 2020). For instance, it is reported that nurses are stressed about a myriad of situations, including worrying about getting infected or inadvertently infecting others and caring for an infectious yet dying patient (Alharbi et al., 2020; Pappa et al., 2020). Moreover, work situations such as erratic and exhaustive work schedules, the lack of personnel protective equipment, and forced deployment to unfamiliar stations are additional burdens. Similarly, they are wary about the social stigma and the uncertainty of whether their employers are genuinely concerned about their welfare (Maben & Bridges, 2020; El-Hage et al., 2020; Zhu et al., 2020).

COVID-19 challenged and brought turmoil to the nurses’ psychological well-being. To mitigate possible physical and psychological damage to the nurses, health facilities advocated the use of mental health services such as psychological first aid, crisis interventions, morale boosters provided by their colleagues, and access to social media and self-help reading materials (Blake et al., 2020; Kang et al., 2020). Interestingly, one study distinctly compared nurses’ feelings and found that those who are less exposed to fever stations appeared to experience more burnout than those on the actual front line. This implies that attention should be provided on an organizational scale, particularly to health and mental wellness interventions (Wu et al., 2020). On the other hand, mounting studies found that nurses who provided direct patient care appeared to be more stressed, overworked, and psychologically disturbed and less fulfilled in their job compared to nurses in other areas of assignment (Zerbini et al., 2020). Hospital nurses, particularly women performing diagnosis, care, treatment, and management of patients with COVID-19, have displayed psychological disturbances such as anxiety, lack of sleep, and depression (Lai et al., 2020).

Researchers have thoroughly discussed the impact of the pandemic on the hospital nurses’ health risks and psychological well-being. However, based on the available literature, there is an evident lack of investigation on the effect of COVID-19 on the nurses’ work outcomes and turnover intention, especially among those deployed in the community. The scarcity of studies in this area prompted the need to explore the nurses’ situation on the ground; hence, this study.

## METHODS

### Research Design

This study employed a descriptive cross-sectional design using self-report questionnaires.

### Participants and Setting

This study targeted 400 nurses to participate; however, only 385 nurses (a response rate of 96.25%) participated. We selected nurse participants who were deployed in community settings under the Human Resource for Health initiative of the Philippine government in response to the COVID-19 pandemic. Particularly, these nurses are assigned as workforce in Municipal Health Offices, Rural Health Units, and Barangay Health Stations. Distinctly, these are nurses who were tagged as the core team in quarantine facilities caring for COVID-19 positive patients who are asymptomatic or who have mild manifestations. Further, these nurses are tasked to monitor PUI (*persons under investigation*) and PUM (*persons under monitoring*) - those who are considered as, respectively, suspected and probable COVID-19 cases - as well as border checkpoints. They operate their stations at an average of 16 hours a day in various municipalities in Samar Province, Philippines.

To reach our target participants, we computed the samples using GPower and discovered that the minimum sample required is 368 at an effect size of 0.05, 0.8 statistical power, and probability level of 0.05 (Scoper, 2015). The researcher collaborated with nurses assigned in the field who are tasked as focal persons-in-charge to conduct Psychological First Aid commissioned by the Department of Health. The core team, who was tasked to provide strategies and interventions to uplift the health workers’ morale on the front line, conducted the collection of data mid-June 2020.

### Instruments

Four self-administered questionnaires were used to gather the data to quantify the variables of this study.

In measuring fear of COVID-19, we used the **Fear of COVID-19 Scale (**Ahorsu et al., 2020). The scale is a valid tool to assess the construct of fear based on its Cronbach’s alpha score of 0.86, suggesting high validity and internal consistency of the scale. The unidimensional 5-point Likert scale is composed of 7 items scored as 1 “strongly disagree” to 5 “strongly agree.”

We also used Schriesheim and Tsui’s (1980) **Job Satisfaction Index** to measure the participants’ satisfaction with their current work assignments. Based on a previous study, the scale demonstrated high reliability and validity coefficients with Cronbach’s alpha of 0.87, implying that the scale is a dependable tool to measure job satisfaction (Labrague et al., 2020). The scale is answerable using a 5-point Likert Scale ranging from 1 “strongly disagree” to 5 “strongly agree.” The job satisfaction scale is composed of 5 items measuring characteristics of work, organizational support, colleagues, salary, and career development, all of which are crucial elements involved in a job.

We then used House and Rizzo’s (1970) **Job Stress Scale** to measure the psychological distress variable. The scale is a valid and reliable instrument based on its high internal consistency of Cronbach’s alpha coefficient score of 0.83. The participant nurses answered this section using a 5-point Likert scale of 1-5 (strongly disagree - strongly agree).

Finally, we used two single-item measures to assess each of the **turnover intention** variables using a 5-point Likert scale ranging from 1 to 5 using the same description as above. Specifically, the organizational turnover intention was assessed with the question, “Given the current situation, I am thinking about leaving this healthcare facility.” Likewise, to assess the participants professional turnover intention, we asked them to rate the statement, “Given the current situation, I am thinking of leaving nursing as a profession.”

### Ethics and Data Gathering Procedure

The authors assured that the study adhered to the basic ethical protocols and was given ethical clearance in one state university with protocol code IRERC EA-0012-I. The researchers secured the approval of the institution and appropriate authorities for the intention to conduct the study. The data gathering was done after the PFA session of the nurses. The participants were informed of the purpose of the study and provided informed consent. It was made clear to the nurses that their participation is voluntary and that they can choose not to complete the questionnaire without any consequence. Further, the participants were informed of their option to remain anonymous and that the data provided was kept confidential.

### Data Analysis

The gathered data were entered into a spreadsheet to facilitate the analysis. The Statistical Package for Social Sciences ver. 23 software program was used in the analysis. Descriptive and inferential statistics were used to analyze the data. To quantify the data, we used frequency counts, percentages, and arithmetic mean. Bivariate analysis and Spearman Rho coefficients were used to assess relationships between fear of COVID-19, nurses’ characteristics, and other key study variables (job satisfaction, psychological distress, and turnover intentions). The predictive analysis between the variables was done using multivariate regression analysis accepted at p < 0.05 level of significance.

## RESULTS

Presented in **Table 1** is the profile of the participants in this study. There was a total of 385 nurse participants with a mean age of 32.65 years old (SD=7.73). Most of the nurses are females (84.2%), married (51.4%), and have completed BS Nursing degrees (96.1%). The nurses have been in active service within the past 7.94 years (SD=7.73) and in full-time service (73.2%) in their present employment.

**Table 1.** Staff, unit, and hospital characteristics (n = 385)

| Characteristics | Categories | Mean | SD |
| --- | --- | --- | --- |
| Age [20-64] |  | 32.65 | 7.73 |
| Year in Nursing Profession |  | 7.94 | 5.79 |
| Year in Present Organization |  | 5.58 | 5.05 |
|  |  | **N** | **%** |
| Gender | Male | 61 | 15.8 |
|  | Female | 324 | 84.2 |
| Marital status | Married | 198 | 51.4 |
|  | Unmarried | 187 | 48.6 |
| Education | BSN | 370 | 96.1 |
|  | MA/MS | 15 | 3.9 |
| Job status | Full-time | 282 | 73.2 |
|  | Part-time | 103 | 26.8 |
| Attendance to COVID-19 training | Yes | 191 | 49.6 |
|  | No | 194 | 50.4 |
| Awareness of existing protocol related to COVID-19 | Yes | 374 | 97.1 |
|  | No | 11 | 2.9 |

**Table 2** presents the key study variables’ mean and standard deviation. Our findings suggest that the participants have a fear of COVID-19 based on the mean score above mid-point (19.92, SD: 5.25). Despite their fears, nurses display a moderate level of job satisfaction (M=3.22, SD=1.09). The high mean scores suggest that there is a presence of psychological distress (M=3.05, SD=0.77), organizational turnover intention (M=2.82, SD=1.21), and professional turnover intention (M=2.87, SD=1.19) among the participants. In terms of health, the participants had a composite mean score of 3.38 (SD=0.87), suggesting they are in adequate health conditions.

**Table 2.** Descriptive statistics of the key study variables.

| **Scale/Subscale** | **N** | **Min** | **Max** | **Mean** | **SD** |
| --- | --- | --- | --- | --- | --- |
| Fear COVID-19 | 385 | 7.00 | 35.00 | 19.92 | 5.25 |
| Job Satisfaction | 385 | 1.00 | 5.00 | 3.22 | 1.09 |
| Psychological Distress | 385 | 1.00 | 5.00 | 3.05 | 0.77 |
| Organizational turnover intention | 385 | 1.00 | 5.00 | 2.82 | 1.21 |
| Professional turnover intention | 385 | 1.00 | 5.00 | 2.87 | 1.19 |
| Health | 385 | 1.00 | 5.00 | 3.38 | 0.87 |

Bivariate analysis was used to examine the correlations between the nurses’ profiles, Fear of COVID-19, and psychological well-being and the nurses’ turnover intentions (**Table 3**). Results revealed that gender is significantly correlated with fear of COVID-19 (t=-2.110, p=0.036), depicting females to be generally more affected by it as compared to males. Moreover, factors such as psychological distress (r=0.354, p=0.001), organizational turnover intentions (r=0.249, p=0.001), and professional turnover intentions (r=0.234, p=0.001) are correlated to the nurses’ fear of COVID-19.

**Table 3.**
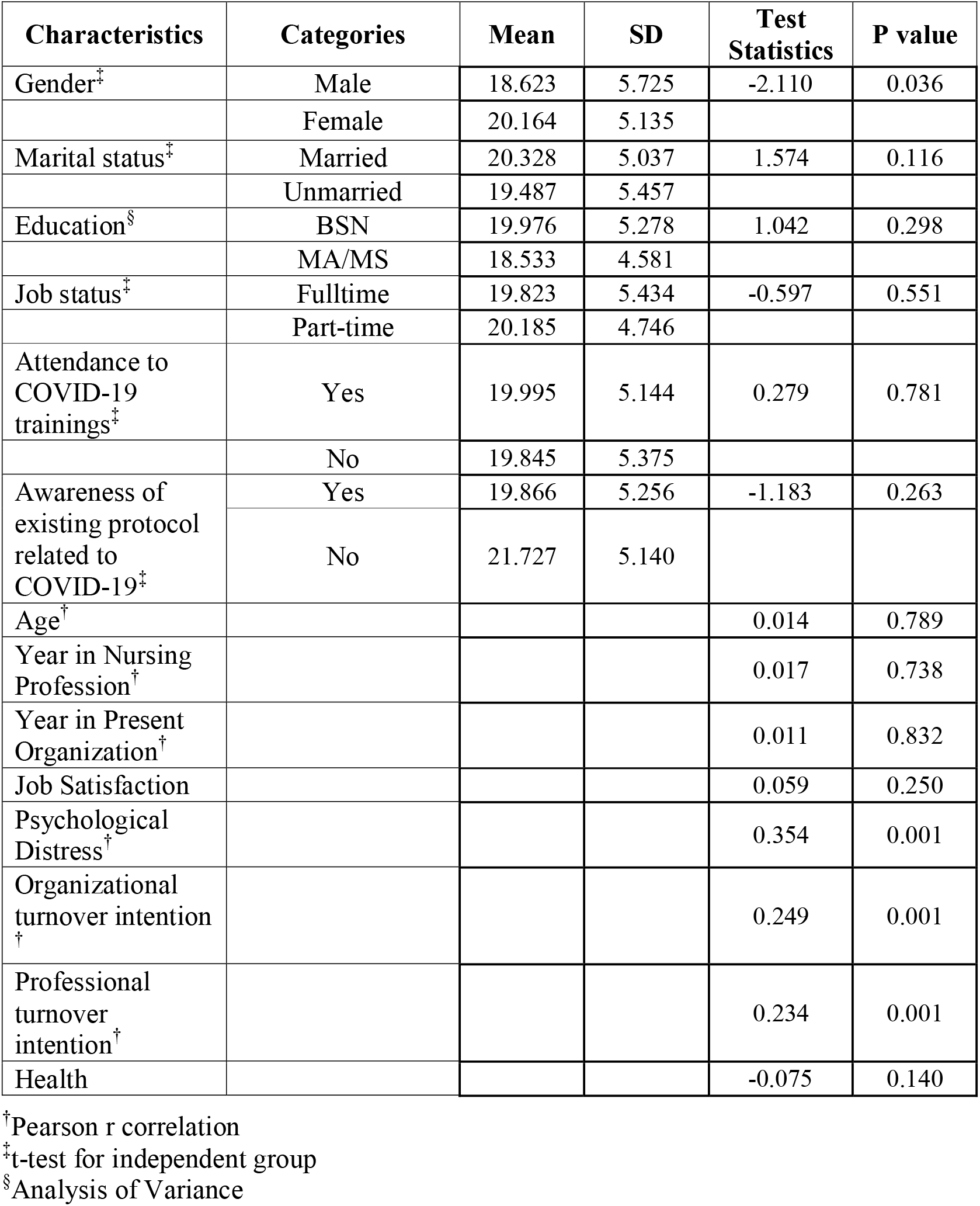
Correlations between key study variables.

**Table 4** presents the multivariate analysis of the key variables of this study. Results showed that fear of COVID-19 influenced the nurses’ psychological distress and turnover intentions (organizational and professional). After controlling the nurses’ profile variables (age, marital status, education, year in nursing, year in the organization, job role, and hospital characteristics including facility size and type of hospital), an increased level of fear of COVID-19 is associated with increased psychological distress (β =0.357; p=0.001), as well as increased organizational (β =0.241, p=0.001) and professional (β =0.221, p=0.001) turnover intentions.

**Table 4.**
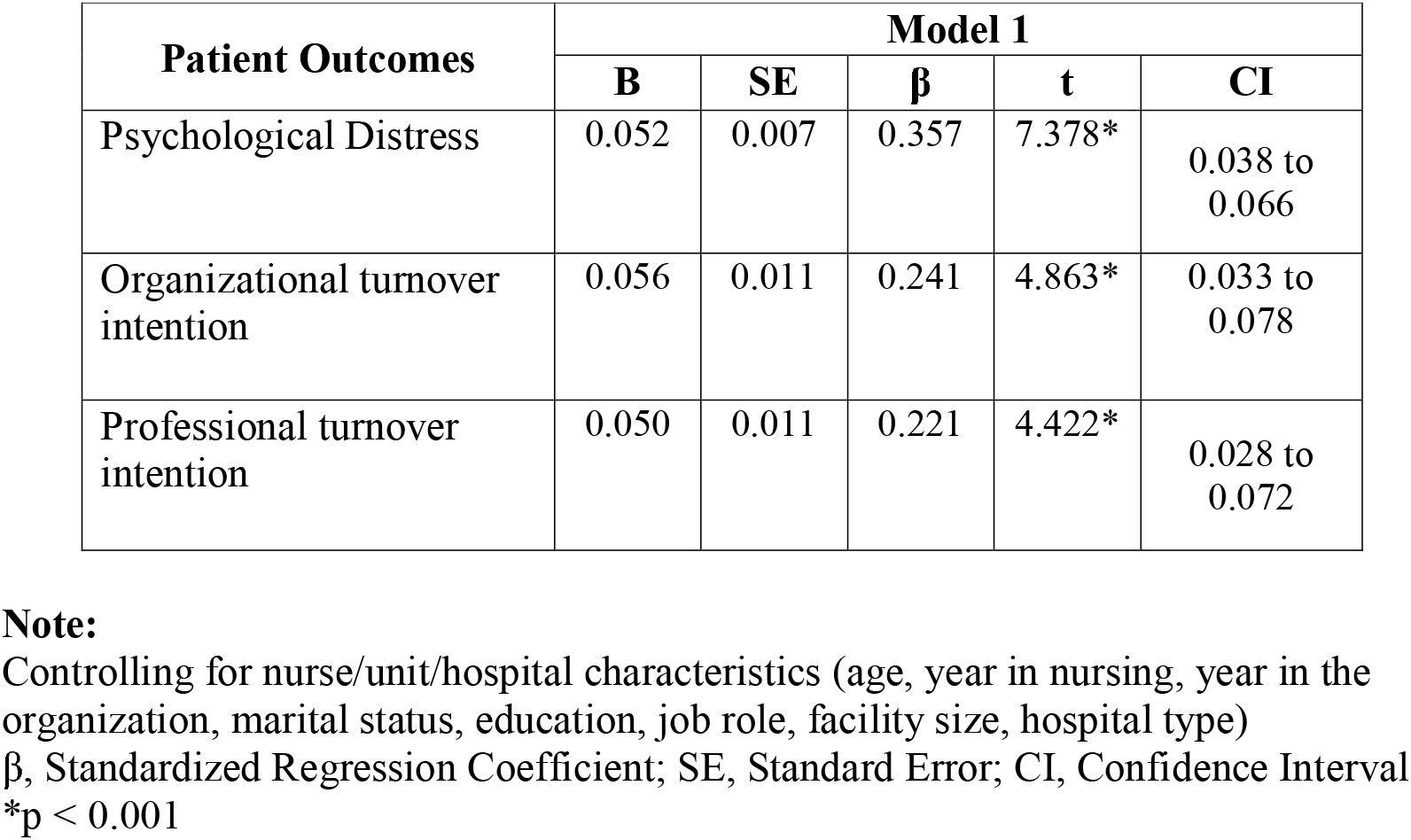
Influence of Fear of COVID-19 on nurse’s job satisfaction, psychological distress, organizational turnover intention, and professional turnover intention.

| Patient Outcomes | Model 1 |  |  |  |  |
| --- | --- | --- | --- | --- | --- |
| | B | SE | $\beta$ | t | CI |
| Psychological Distress | 0.052 | 0.007 | 0.357 | 7.378* | 0.038 to 0.066 |
| Organizational turnover intention | 0.056 | 0.011 | 0.241 | 4.863* | 0.033 to 0.078 |
| Professional turnover intention | 0.050 | 0.011 | 0.221 | 4.422* | 0.028 to 0.072 |

**Note:**
Controlling for nurse/unit/hospital characteristics (age, year in nursing, year in the organization, marital status, education, job role, facility size, hospital type)
$\beta$ , Standardized Regression Coefficient; SE, Standard Error; CI, Confidence Interval
\* $p < 0.001$

## DISCUSSION

The authors believe that this is the first study to investigate the welfare of nurses deployed in a community setting. This study’s primary goal was to assess the fear of COVID-19 and its influence on the psychological well-being and turnover intentions among nurses assigned to care and manage COVID-19 patients outside the hospital setting. Our results revealed that community nurses are relatively young and have been in government service since the onset of their professional careers. The unparalleled health crisis caused by this pandemic created a conundrum of anxiety and psychological distress, especially among nurses, in both hospital and community settings.

The COVID-19 pandemic caused a great deal of fear for nurses in various countries, including China (Hu et al., 2020), Taiwan (Feng et al., 2020), Italy (Bagnasco et al., 2020), Singapore and India (Chew et al., 2020), and, in particular, the community nurses in the Philippines. Nurses in the field display moderate fear of COVID-19 even when caring for asymptomatic to mild cases. This suggests that fear of COVID-19 is universal to all nurses. Studies have also presented that hospital nurses reported to be afraid mostly of the fear of transmission and the consequences of inflicting it on their patients (Apisarnthanarak et al., 2020; Sun et al., 2020). This supports the report that all health workers, especially nurses at the forefront of this difficult time, are challenged regardless of their institutional setting (Jackson et al., 2020; Boyra & Legros, 2020). The rapidly increasing number of COVID-19 cases from local returnees coming from endemic areas, as well as the regularly changing quarantine protocols, lack of established health information, longer work hours, laborious contact tracing, and depleted supplies of personal protective equipment, physically and emotionally exhaust community nurses, increasing their fear of contracting COVID-19. To a greater extent, these nurses in the Philippines also share the burden with other human resource workers in a multi-sectoral approach to control community outbreaks in the country (HCT Philippines OCHA, 2020).

Remarkably, our findings suggest that females are more fearful of this pandemic than male nurses. This finding is similar to other studies reporting a correlation between the female gender and the fear of COVID-19 (Lai et al., 2020; Mazza et al., 2020). In addition, our finding is related to reports which found that female nurses tend to be extra cautious with infection control practices, especially when caring for patients with infectious disease, because of perceived vulnerability to infection and as a precaution to avoid infecting their families (Efstathiou et al., 2011; Jackson et al., 2020; Russell et al., 2018).

Similarly, nurses deployed in the community reported high psychological distress. The scarcity of literature on nurses in the community setting in the time of COVID-19 hinders us from comparing our findings. Nevertheless, psychological distress is common among nurses treating cases of COVID-19, such that in the case of community nurses in this study, anxiety, fear of getting infected, and social isolation are the dominant psychological stressors. Meanwhile, nurses in a hospital setting are distraught psychologically and manifest symptoms such as loss of appetite, sleep disturbances, nervousness, and suicidal ideation, especially in critical care units (Shen et al., 2020). Also, there is a higher perception of perceived stress and depression observed among those working in respiratory medicine departments (Ma et al., 2020). On the other hand, emergency nurses exhibit depressive symptoms and Post Traumatic Stress Disorder (Song et al., 2020). Moreover, one study found that psychological distress is evident to both ordinary nurses and advanced practice providers (Shechter et al., 2020). This suggests that regardless of their workstations or expertise, nurses are vulnerable to the psychological implications of COVID-19.

Our results also revealed that nurses in a community setting show a moderate level of job satisfaction, perhaps because they are less stressed than nurses in hospitals’ acute and critical care areas specializing in COVID-19 cases. Compared with nurses in community settings, hospital nurses encounter more stressful work situations because of the high demand for nursing care. The literature revealed that workload is the primary stressor that influences nurses’ job satisfaction (Bautista et al., 2020; Lu et al., 2019). In the case of the community nurses in this study, the moderate level of job satisfaction may be attributed to their perceived better health conditions. This is similar to the study of Zhang et al. (2020) in Iran, who found that better physical health conditions influence job satisfaction in healthcare personnel despite the odds of a risky work environment.

Additionally, there is job satisfaction among nurses in the community because they can function in their nursing roles, especially in these unconventional times. Nurses choose to face the adversities of COVID-19 because they can personify the value of altruism and teamwork attached to their jobs as care providers (Sun et al., 2020). The literature revealed that nurses tend to be work-excited and highly committed to their work, especially when they are professionally challenged (Chang et al., 2020). Nurses display more dedication to their professional duties at their expense in times of pandemic (Fernandez et al., 2020), as projected by these nurses in the community.

Moreover, our results suggest that despite their job satisfaction, community nurses are not married to their jobs during this pandemic. Our findings indicate that these nurses would still prefer to leave their jobs and shift to another career. This contrasts with other studies that discussed how job satisfaction negatively influences turnover intention (Labrague et al., 2020; Li et al., 2019). Our findings are similar to Jung et al. (2020), who found high turnover intentions among nurses caring for patients in infectious disease outbreaks. COVID-19, as a highly contagious and pathogenic disease, has taken the lives of thousands, and healthcare providers could be discouraged by the risk to their lives. While it is true that the need for nurses at these times has never been higher, the novelty and the enormous ramifications of COVID-19 force these nurses to consider leaving their jobs and shifting to another means of living.

Finally, the regression analyses in this study revealed that the nurses’ fear of COVID-19 is related to their psychologically distressed disposition and turnover intentions. This finding is similar to the mounting studies associating COVID-19 to nurses’ psychological distress (Chew et al., 2020; Schecter et al., 2020; Fernandez et al., 2020). However, as of this writing, there is no literature discussing the influence of fear of COVID-19 on nurses’ organizational and professional turnover intentions. Nonetheless, fear of COVID-19 has brought enormous stress and psychological distress to the nurses, influencing their high turnover intentions. The findings of this study relate to previous studies discussing how burnout, moral distress (Hatamizadeh et al., 2020), job demands (Boudrias et al., 2020), and work pressure influence turnover intention and work engagement among nurses in the community (Li et al., 2019).

The pandemic has undeniably challenged the roles of nurses. The real scenario at present is that nurses, including those in community settings, are placed in a dilemma of whether to protect and save patients or preserve themselves for their families and loved ones who are also depending on them. The vulnerability of life outweighs the call of duty in these desperate times, considering that nurses, too, have families and loved ones waiting for their safe return from the battlefield with this unseen yet deadly virus. Despite this threat and the psychological distress, nurses continue to function and fulfill their professional tasks in caring deserved by their patients and communities.

### Implications to Nursing Practice

Community nurses facing persons who may be COVID-19 positive when doing contact tracing and specimen collection are facing a risk on their end, which may be attributed to their anxiety or fear of viral transmission. Consequently, these nurses need attention and aid from nurse administrators and other executives in the health sector. To provide nurses with assurance, comfort, and mental health, there is a need to increase capacity building to boost issues on caring and patient management incapacity, institute psychological support services, and stress reduction. For instance, the provision of enough PPE and testing for COVID-19 exposure is one strategy that can be followed to mitigate feelings of fear and anxiety among these nurses (Zhang et al., 2020). Studies also suggest physical exercise can help (Schecter et al., 2020). However, these strategies should be initiated by colleagues outside the COVID-19 areas to drive more energy and enthusiasm to these already psychologically disturbed nurses. There is a need for a collective effort to provide all the support possible to increase the psychological well-being of nurses on the front line. Additionally, providing their basic needs of health safety during isolation and quarantine and implementing more rest and relaxation activities are strategies that organizations can consider to reduce the worries, anxieties, and fears of these nurses. In this time where social isolation is the new normal, online support through tele-health is imperative to provide psychological support to these nurses, especially when face-to face is not possible (Zaka et al., 2020; Zhou et al., 2020). These are just few of the strategies currently being implemented by organizations in different countries. Ultimately, it is also a priority to ascertain and explore the real causes of fear and psychological distress to address these issues better and avoid turnover intention.

## LIMITATIONS OF THE STUDY

This study is not without limitations. First the design used a cross-sectional approach and therefore cannot establish causality. Second, we gathered participants in one province in the country; hence, the study does not intend to establish generalizability. The construct of fear and psychological distress is better assessed using a qualitative design to provide a more in-depth understanding of the nurses’ experience in the light of COVID-19.

## CONCLUSION

This study revealed that community nurses share the same experience of fear of COVID-19, similar to nurses working in a hospital setting, with female nurses appearing to be more fearful than male nurses. Further, we conclude that with increased fear of COVID-19, the nurses’ psychological distress, as well as their organizational and professional turnover intentions, increases. Nurse leaders and organizations must assess these nurses’ needs and initiate measures to provide the necessary psychological support to these nurses on the front line.

## Data Availability

The data will be made available upon request

